# Management of conductive deafness from Otitis Media with Effusion (known as ‘glue ear’) in children using bone conduction headsets when grommet operations were unavailable during COVID-19

**DOI:** 10.1101/2021.01.21.21249496

**Authors:** T Holland Brown, Isobel Fitzgerlad O’Connor, Jessica Bewick, Colin Morley

## Abstract

**Background:** Otitis Media with Effusion (OME) causing hearing impairments affects ∼1 in 10 children starting school in UK/ Europe. 80% have at least one episode with most having conductive hearing loss. Studies showed children with OME hear better with bone conducting headsets. During COVID-19 we investigated whether children with deafness secondary OME, without access to audiology or grommet surgery, could be aided with bone conduction kits and the HearGlueEar app.

**Methods:** Starting July 2020, during COVID-19, children aged 3-11 years with OME and on a grommet waiting list were invited to a single arm, prospective study. They received the kit, instructions and HearGlueEar app by post. By 3 weeks parents were asked to charge and pair the devices, attend a remote consultation and complete an OMQ-14 questionnaire. Remote follow-up lasted 3 months. Outcomes: ability to use the equipment, complete the questionnaire about child’s hearing and behaviour before and with the equipment, declining grommet surgery or where deafness resolved, and give opinion about the intervention.

**Findings:** 26 children enrolled. Families used the kit at home and school. Most found remote consultations positive and convenient. OMQ-14 responses were 90% positive. Comments were: “Other people have said, wow his speech is clearer.”, “It is making a real difference at home.”, “He said over and over again, “I can hear everybody, wow, wow, wow.”, “It is no exaggeration to say this has made an astronomical improvement to his quality of life”. One child reported “I can hear my best friend again”. “She is getting on really well with the headphones - pairing them with the iPad at home is simply brilliant.” Three families continued with the headset to avoid grommets.

**Interpretation:** Posting a bone conduction kit, HearGlueEar app and remote consultation is effective support for children with deafness secondary to OME.

**Funding:** None

## Background

Otitis Media with Effusion (OME) is the commonest cause of childhood hearing impairment and can recur for long periods.^1,2^ It affects about 1 in 10 children starting school in UK/ Europe. 80% have at least one episode, with most having an associated conductive hearing loss. Conductive hearing loss in children is 100 times commoner than permanent sensorineural hearing loss.

OME tends to cause a mild to moderate, unilateral or bilateral, conductive hearing loss that fluctuates. It may resolve or persist. Children with OME struggle to hear, such as listening to a teacher, particularly with background noise. The group most affected are under 8 years old, which is a critical time for development, speech acquisition, learning reading, writing, spelling and phonics.^3^ Deafness at this time interferes with speech development, language, communication, auditory processing, self-esteem, socialisation, listening and learning.^4^

The NICE guideline, “Otitis media with effusion in under 12s: surgery” suggests children with OME should have a period of “active observation” to see if OME resolves, before offering a grommet operation or hearing aid, and parents, “might like to ask” professionals, “Is there anything I can do to help my child while we wait”? ^5^ Practically this means wait and see if hearing impairment resolves. During COVID-19 local community paediatric audiology services closed to referrals between April and September 2020, and later opened to a large waiting list. Hospital audiology services also were reduced and had long waiting times. Grommet operations were in the lowest priority for elective surgery in the UK.

Our previous studies investigated aiding the hearing of children with OME using bone conducting headsets where sound vibrations pass through skull to the cochlea, bypassing the middle ear. The volume of sound delivered with these headsets needs little adjustment. Parents (with normal hearing) were told to try it themselves and if necessary, adjust the volume to ensure it wasn’t too loud or quiet. As the headsets do not cover the ear, near-by sounds may be heard. Studies showed children with OME heard speech at a quieter decibel level wearing the headsets. ^4^ The majority also used the HearGlueEar app (with songs, games and audiobooks created to support children with OME) as a resource and found it helpful.^6^ Even children without OME showed improved word recognition when wearing a headset.^7^ In previous studies, doctors and audiologists introduced and explained use of bone conducting headsets face to face.

The aim of this study was to investigate if children with conductive hearing impairment secondary to OME, who had no access to audiology or surgery during COVID-19, could be helped remotely without face-to-face professional support, using an affordable Raspberry Pi Bone Conduction Headset (Raspberry Pi, Cambridge, UK) and Microphone sourced for this study (hereafter called kit) simple written and video instructions, and the HearGlueEar app sent by post.

## Methods

This study started in July 2020 during the Covid-19 pandemic. Children diagnosed with OME (hearing worse than 25dB at 2 or more frequencies in at least one ear) and those on a grommet operation list were invited to enrol in a single arm, prospective, study assessing the use kit. If parents agreed, they were posted the kit for use at home and school with instructions and details of the HearGlueEar app to use with it if they wanted to. (https://hearglueear.wordpress.com/). The parents were told to follow instructions and asked to self-check the headset volume (if parent had normal hearing) to a comfortable level, before handing it to their child.

Within the first three weeks, parents answered questions about their child’s hearing by completing a OMQ-14 questionnaire with and without the kit.^8^ Follow-up consultations were remote, by video or telephone over 3 months. The study was registered IRAS 262816 and approved by the Solihull REC and Health Research Authority (HRA).

### Patient and public involvement (PPI)

PPI, funded by the East of England Research Design Service, as well as feedback from patients involved in previous local OME studies, captured patient concerns, importance of empowering families and assisting self management through the OME pathway, which directed the focus of this research study. A website (www.hearglueear.wordpress.com), initially made solely for research participants, communicated study details, results and publications.

## Results

Twenty-three (82%) were recruited from the local grommet (ventilation tubes) operation list of 28 children. Two dropped out due to normalisation of hearing, their parents were reluctant to return the equipment. One re-joined the study when OME returned. Four participants were recruited from paediatric audiology clinics when referred to ENT. 26 children between 3-11 years (average 5 years) were enrolled. One had a cleft palate, one atypical Turner syndrome and one social, communication and learning difficulties. The average hearing loss at frequencies of 0.5, 1, 2, 4 kHz ranged from 0-65 dB before lockdown in March 2020. The mean for each frequency was 35, 30, 25, 30 dB in the right ear and 30, 27, 20, 27 dB in the left ear. This is a mild hearing loss. For bone conduction it was 6, 7, 9, 4 dB at 0.5, 1, 2, and 4 kHz respectively.

14 children had hearing difficulties ranging from 9 months to 8 years (average 2.9 years) before the study. 89% (23) described speech/language difficulties or delay. 58% (15) were considered academically behind their peers, mostly with phonics and reading, compared with 2.7-14.6% nationally.^9^ 35% (9) had behaviour difficulties compared with a similar population with 3.2% of girls and 6.7% of boys.^10^ 31% (8) had attention problems, 42% (11) anxiety or emotional concerns where the national average is ∼ 4% ^10^ 15% (4) friendship or social communication concerns, compared with around 1% in the UK.

At the three-week telephone consultation, no family required help to use the kit. Twenty started using it before the consultation. Five had not used it: 1 arrived the morning of the consultation; 2 had no microphone, 2 waited for researcher’s phone call. No one wanted additional guidance. All paired the device using written instructions. Seven (28%) used additional video instructions. As the study progressed, to follow-up at three months 80% (21) wanted a telephone consultation and 20% (5) a video consultation.

### How families used the kit and their comments

Table1. shows parent’s opinions on their child’s hearing and the effect of the kit. It shows that before the use of the kit, their hearing was generally poor, they often misheard words and in a group, they often had difficulty hearing. With a kit their hearing was better, they rarely misheard words and in a group listening was easier. Some reported their child to be less anxious.

**Table 1.** Parent’s reported opinions on their child’s hearing. These were the relevant questions to the study extracted from the OMQ-14.

|  | **During 3 months before the study**<br>**(April, May June, or July) during**<br>**school 'lockdown'** |  | **Used Kit for <3 weeks** |  |
| --- | --- | --- | --- | --- |
| Describe child's hearing | Normal | 0 | Normal (with headset) | 23 |
|  | Slightly below normal | 6 | Slightly below normal | 1 |
|  | Poor | 12 | Poor | 0 |
|  | Very poor | 7 | V poor | 0 |
|  | Unsure | 1 | Unsure | 2 |
| Has child misheard words<br>when not looking at you? | No | 0 | No (or "improved") | 17 |
|  | Rarely | 2 | Rarely | 5 |
|  | Often/ Always | 23 | Often/ Always | 1 |
|  | Unsure | 1 | Unsure | 1 |
| Has child had difficulty hear-<br>ing when with a group? | No | 0 | No | 15 |
|  | Rarely | 2 | Rarely | 7 |
|  | Often | 11 | Often | 1 |
|  | Always | 12 | Always | 0 |
|  | Not sure | 0 | Unsure (Only used headset paired to<br>computer/ tablet) | 2 |

One family said, “We were so desperate, and we had run out of options. This is just brilliant. We are over the moon to be part of the study”. “I have been wanting to support [my child] with something like this”.

Four had trouble pairing or operating the equipment at some point during the study. Four different families were reminded the kit could be paired to other equipment. Therefore, the role of the researcher to trouble shoot intermittently was important.

Families used the kit in different ways. Most families tried all options. Some preferred using the headset just paired to the microphone, some used the headset exclusively paired to phones/iPads (figure3). Some used the HearGlueEar app regularly, others did not. 66% (17) used the app at home, 8% (2) on car journeys and 26% (7) did not use it at all. Some used the headset paired with a tablet for Zoom calls with family, health professionals, music teachers, school/online lessons or audio-books and films.

All the children looked after the kit. No one lost or broke the headphones. Two parents reported their children were, “very protective over their headphones”. Three families reported their child, “had worked out how to pair it with everything in the house”.

Some of the parents’ comments when their child first used the kit: “He put his headset on yesterday, and his face lit up like I’ve NEVER seen! He said over and over again, “I can hear everybody, I can hear everybody! Wow, wow, wow, I’m freaking out!”, “It is no exaggeration to say this has made an astronomical improvement to his quality of life”, “I wish I had filmed it when he first tried it on, he said, “I can actually hear”, “The look on his face when he could hear was priceless”, “He said ‘ohmigod! I can hear everything’. It was obvious by his face. He was so surprised”. One child reported, “I can hear my best friend again” and “thank you for giving it to me”, “The TV volume stays down now, we just place the microphone by the TV”, “The interaction between the boys [siblings] has been better”, “She [10 y] is relying on it enormously…. has paired it to everything… It is now the way she watches the TV”, “She is getting on really well with the headphones - pairing them with the iPad at home is simply brilliant. She loves the freedom they give her and obviously, the hearing ability.”, “There have been fewer arguments because he could communicate more effectively”.

### When children returned to school

When school restarted families had to decide whether the child should ask to use the kit at school. None were told they must take it to school. Four families (15%) said their child needing more hearing support as they started the new school year: Two were seated in poor listening positions at the back of the class and one said there was a lack of teaching support for their child!s hearing needs, and one had been placed in a different room without friends who understood her speech.

All children used the kit at home, 58% (15) took the kit to school or nursery. One wanted to take the kit to school but parents advised against it. One teacher said she, “would see” if the child needed it in class. The child continued to use it at home.

Eleven (42%) prioritised the use of the kit outside school e.g. shopping, bike-rides, socialising, online learning.

Some comments from the parents were: “His speech is coming on so well in just a week”; “Her vocabulary is also markedly improved, as is her reading”; “According to his teacher he is now sitting for circle time for the whole amount of time, before he would get up halfway through.”

Average hearing levels before the study were similar between those who took the kit to school and those using it at home.

Families used the teacher information sheet when taking the kit into school. One family commented that the school took their child’s hearing more seriously as a result of the need for the headset: One parent commented the headset was a visual cue to others their child needed support. Three parents mentioned schools needed more support, or more resources, to use the kit at school.

Three parents contacted their school Special Educational Needs Coordinator. This had advantages and the child was more likely to use the kit seamlessly at school. In two cases the teacher addressed the class about the headset. Both these helped the child to talk about their hearing and the headset with their peers and both children became active users of the headsets. Parents of one child explained that he checked his bag every day before school to see the kit had been charged overnight and placed back in his bag.

Eight schools or nurseries followed infection control advice to use antiseptic wipes when transferring equipment between home and school. However, seven wanted different equipment at home and school. Of these, two schools only needed replacement microphones as that was the only shared part of the device. Five wanted full kit in school. One quarantined the kit for two days before allowing them to be used (this was not supported by any written guidelines.) Two who had school-use-only equipment, allocated it to be used by only one teacher, which meant children described at follow up how they did not have access to their equipment with substitute teachers, or subject-specific teachers e.g. physical education (PE). This led to children trying to decide which lessons they needed it for most. One child said PE was the worst listening place, but she needed the kit for normal classes.

By the time of writing (Dec 2020) eight children had repeat audiometry assessments which showed four had a marginally worse audiogram than before lockdown, one a similar audiogram and three an improved audiogram.

### Follow up by three months

Prior to the pandemic, one child who had struggled with noise hypersensitivity as well as OME, had coped with his anxiety around this by becoming dependent on noise cancelling headphones at school (often in the playground). During the study his use of ear defenders was replaced by the kit because hearing loud noises was less surprising or worrying. His mother reported that when the “School had a fire drill, he didn’t scream and cry or ask for his ear defenders”. Noise cancelling headphones are more expensive than the kit. Using the kit for children with OME and noise hyper-sensitivity is a future research area.

At the end of the study, families were able to keep the kit and contact the team if they had questions or concerns, in line with ethical approval. Three families said they would rather continue with the headset and avoid a grommet operation. No child had a grommet operation by Dec 2020 (9 months after lockdown). Three had the kit until their glue ear resolved. This is six avoided grommet operations (23%) by this time. Cost of the kit will be ∼£50 and 23% of grommet operations were avoided suggesting there could be a cost savings to the NHS. Further follow-up is anticipated.

Further comments from the parents: “It is making a real difference at home”, “I can’t tell you how much it has helped”, “It has made a massive difference to her.”, “It’s nice to get something to help”. Sixty-six percent of families, with a sibling who also had glue ear, asked for another kit.

## Discussion

Families wanted to help their child after a long period of hearing loss and so it is not surprising, they were positive about this intervention.

An important finding from this study was that most families set up kit without face-to-face professional help using written instructions, and a short video demonstration. Four had trouble with pairing or operating the equipment but this was rectified remotely. One family who could use the kit at home found the teacher struggled to pair it with the microphone. We realised the teacher’s instructions were brief and a more comprehensive information pack for teachers would help. One family had a challenging home environment and so the teacher took charge of the headset and operated it successfully at school. For the kit to be used successfully it is important for a professional to be available to answer questions and have spare equipment available.

Remote management of glue ear in this way has many advantages: improving children’s hearing at an important stage of their development when there is little alternative help, preserving face-to face hospital appointments with surgeons and audiologists for those needing them most, reduces travelling to an audiology or hospital clinic with small children, empowers the parents to support their children and saves costs for the NHS.

Importantly the parents’ comments showed their children benefited from the kit, helping them hear better at home and school, and in some cases improved their pronunciation, behaviour and socialisation.

Despite the fact that glue ear often fluctuates from a unilateral to a bilateral loss and with time, grommet operations are often only offered if there is evidence of audiograms indicating a bilateral loss. The simple intervention used in this study can help their hearing while waiting for possible surgery or resolution of the deafness.

This study highlights that parents wanted more support. Some explained that when their child’s glue ear and deafness fluctuated, but the hearing test was normal, they were discharged from the clinic without a plan if it recurred. Care plans are used in other paediatric specialities where recurrence of a condition is a possibility such as epilepsy or asthma. An example care plan is in the appendix.

Some parents preferred to continue to support their child’s hearing with this kit rather than have a grommet operation. This would be a cost saving to the NHS. In line with ethical approval, patients were able to keep the equipment at the end of the study.

It could be argued that hearing services have a responsibility to offer affordable easy and effective hearing support to children with OME, if they have to wait for, or do not have access to, grommet operations.

None of the children had teacher of the deaf (TOD) support at the outset, but due to one child coming into contact with a TOD and the TOD acknowledging that because the child had a hearing loss supported with hearing equipment in school, the criteria for TOD involvement was fulfilled and so received support at school. This led to a discussion about the other children in the study with TOD’s, and subsequent TOD support for some. Families chose whether their child should take the headphones to school or not. Just over half took it to school. Although no child started with TOD involvement some were subsequently helped in school.

Future research should investigate hearing support for children with Down syndrome as they are prone to glue ear.^11^

A weakness of the study is that it was opportunist, short term, not randomised and could only recruit a relatively small number of children. Despite this, it provided the opportunity to show that children with conductive deafness could be simply and effectively supported.

Using the HearGlueEar app, “How’s my hearing” section (with two hearing screening options: one based on tones and the other on hearing speech), parents were able to self-manage the OME with the kit.

During COVID-19 children needing to hear in new listening situations such as zoom calls, on-line class learning/ teaching found the kit helpful. Hearing in these situations needs addressing from now on. Social distancing and use of face-coverings further hindered hearing and the kit helped mitigate this.

As the NHS backlog grows during the pandemic, innovations are needed to overcome the prolonged wait for grommet insertion which is considered a low priority operation. As a low cost, low risk, intervention this study shows that even those with a mild hearing loss benefitted from a bone conducting headset.

## Conclusion

Posting an affordable bone conduction kit, information about how to use it, and details of the HearGlueEar app to families is a simple, effective way to support children with conductive hearing loss secondary to glue ear, or for children waiting for a grommet operation. Families found this was acceptable and a positive way of helping their child’s hearing.

### Summary box

#### Key points

- New findings from this study show families who have a child with OME can be sent kit through the post and be managed remotely.
- Children and parents benefitted from being empowered to self manage OME.
- Children with hearing loss had new listening challenges during the pandemic: Social distancing affecting signal-to-noise ratios; face-masks obscuring lip-reading; listening to education classes/ music lessons/ socialising on-line. The kit (bone conduction headset; microphone; HearGlueEar app) used, helped children overcome these challenges. Children used the kit in other situations too such as when shopping, on bike rides, in the classroom (as schools re-opened).
- The impact of this study on future health care : OME can be managed remotely, this approach decreases the number of grommet operations parents opted for and impacts/ improves the child’s reported hearing as well as their access to learning and education at home or at school.

### Research in context

#### Evidence before this study

OME causing hearing impairments affects 1 in 10 children starting school in UK/ Europe. 80% have at least one episode with most having conductive hearing loss. It is 100 times commoner than sensorineural deafness. They often have developmental, educational, speech, language, cognitive and behavioural sequelae and need timely intervention to reduce these effects. They rarely receive hearing support while waiting to see if they need a grommet operation. Standard hearing aids are not ideal due to the fluctuating hearing loss. Bone conducting headsets have been shown to improve their hearing.

#### Added value of this study

During COVID-19 pandemic, children with OME were managed with affordable bone conduction headset kits without professional support. More than half used the kits at school and home. Quality of life measures improved. Children also used the kit to assist in new listening challenges such as zoom calls or remote education as well as social distancing and masks preventing lip reading. Some declined grommet surgery or the OME resolved.

#### Implications for practice

This affordable, remote approach, could provide hearing support with a bone conduction and microphone kit to children with conductive hearing loss from OME, improve quality of life for children, reduce the grommet operation and audiology waiting lists and save costs for the NHS.

**Figure 1.**
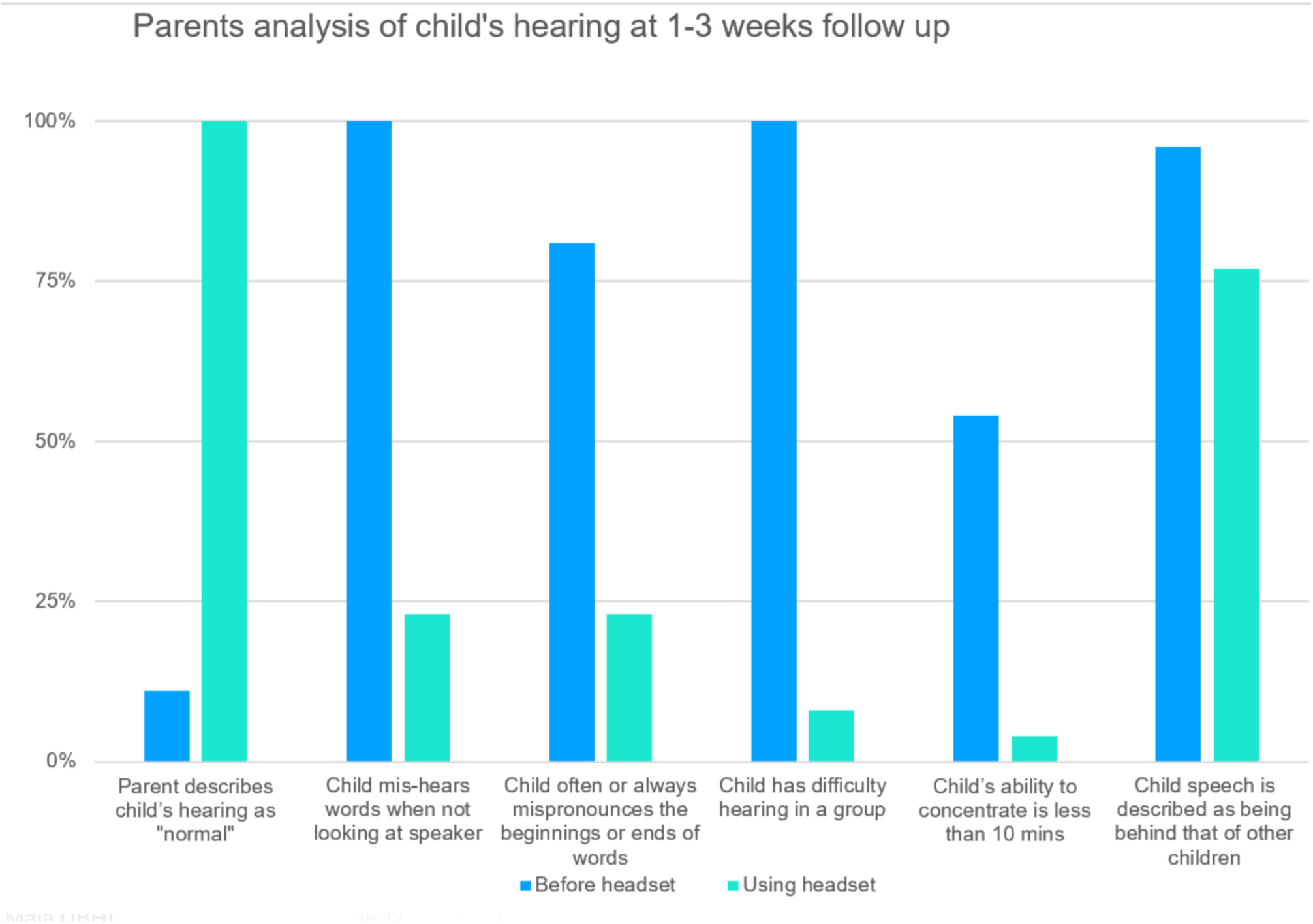
Parents’ analysis of child’s hearing at 1-3 weeks follow-up

**Figure 2.**
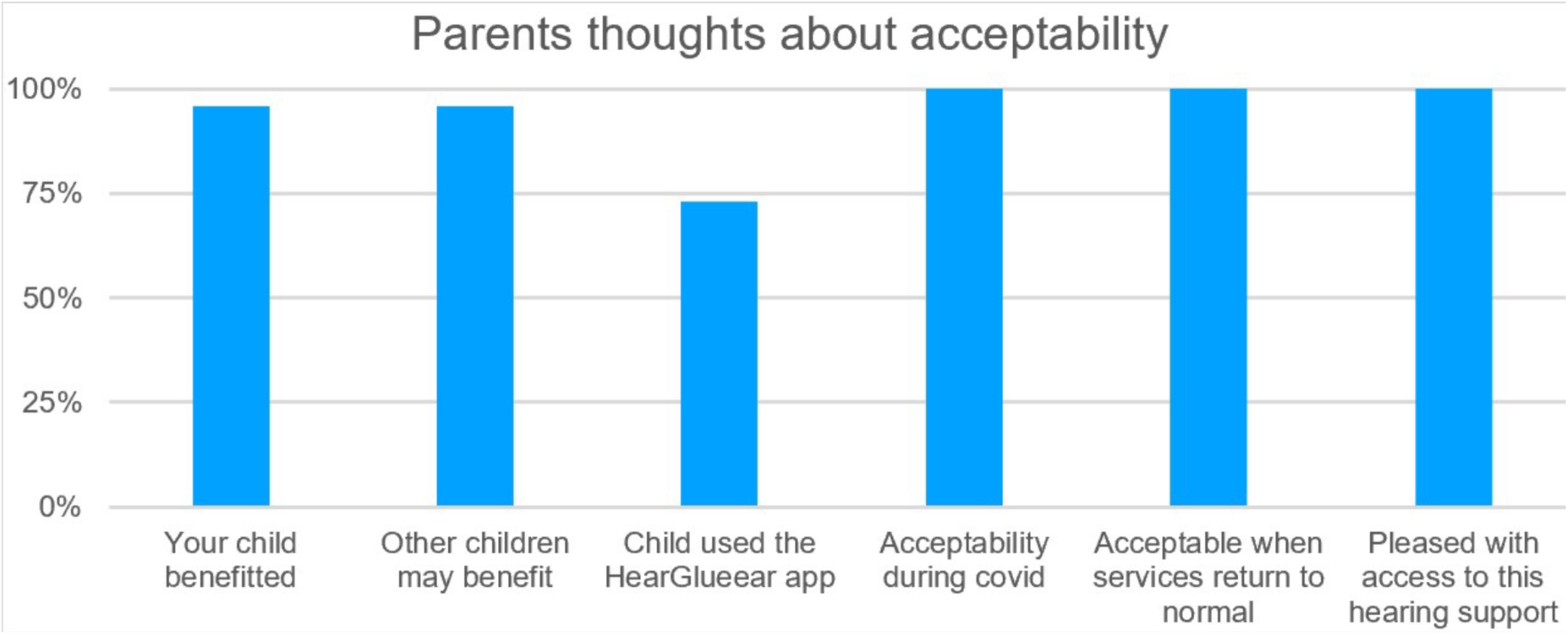
Parents responses about acceptability of the bone conduction kit.

## Data Availability

Data sharing of anonymised patient data collected for the study can be made available to others. Other details of the study have been published on https://hearglueear.wordpress.com/

https://hearglueear.wordpress.com/research-during-covid-for-children-with-glue-ear-who-have-been-considered-for-a-grommet-operation/

## Appendices

### 1. Comments parents wrote about their child using the headset, sorted into categories

1. Comments parents wrote about their child using the headset, sorted into categories.
| Categories | Positive comments | Neutral comments | Negative comments |
| --- | --- | --- | --- |
| **Impact on hearing** | "He says to everyone, "I can actually hear you now" “.<br>"He put his headset on yesterday, and his face lit up like I've NEVER seen! He said over and over again, "I can hear everybody, I can hear everybody! Wow, wow, wow, I'm freaking out!". It is no exaggeration to say this has made an astronomical improvement to his quality of life"<br>"I wish I had filmed it when he first tried it on, he said "I can actually hear""<br>"The look on his face when he could hear was priceless"<br>"He said "ohmigod! I can hear everything." It was obvious by his face. He was so surprised".<br>**Child:** "It definitely helps with being able to hear".<br>"I can hear my best friend better now" | "He did like them. They are comfortable. He said he could hear better, that's the only thing I can go off."<br>"We only have them connected to the iPad so I can't tell if she can hear better" |  |
| **Impact on anxiety/ worry** | "Definitely happier and calmer when wearing headphones".<br>"She is less emotional, a happier child when she has it on and less tired from lip reading".<br>"School has a fire drill, he didn't scream and cry or ask for his ear defenders" (for a child who had previously become dependent on wearing noise cancelling ear defenders for anxiety around loud sounds at school)<br>"He is happier when he is wearing them" | "He's more chilled, he didn't get shouted at". | "She feels anxious when headphones aren't working or charged". |
| **Impact on speech and language** | "She speaks more normally [when wearing the headphones], phonics at school - she is pronouncing 'ch' and 'm' clearer".<br>"He used to get the next door neighbour's name wrong but when he said it right the other day, we was all shocked, even she was shocked".<br>"She wears the headphones to sing along to Mamma Mia and she now sings the words more clearly".<br>"It's made a big difference to how she hears and her speech. She had gone back [during lockdown] to signing for 'hungry' and 'drink' from Mr Tumble. The signing stopped with the headphones".<br>"He was so upset when it [headphones] wasn't working. Little things started creeping in. His speech regressed really quickly. He wasn't starting his words properly".<br>"Better pronunciation when wearing it".<br>"It's worked really well for her, aiding her speech development. It's a positive outcome for her".<br>"He speaks at a better volume level himself when he is wearing headphones".<br>"His speech is coming on so well in just a week".<br>"There is a quite a change in how he is saying words. He is able to correct himself more easily when wearing the headphones".<br>"Other people have said, "wow his speech is more clearer" | "Her speech is generally improving, although I'm hesitant to say the extent etc. I've been surprised at times the length of word she's used or the complexity of the sentence or that she's heard and repeated something that I've said. However, there are still lots of times when her speech is slurred, or I cannot immediately get what she is trying to say and it's not clear." | "His speech is still bad whether he wears them or not". |
| **Impact on school/ learning** | "His learning is much quicker ..it's given him a kick start".<br>"Her vocabulary is also markedly improved, as is her reading".<br>"According to his teacher he is now sitting for circle time for the whole amount of time, before he would get up half way through." |  | "She doesn't want to wear them much at school as she is conscious of what the other children in her class think of her." |
| **Teachers** | "He engages with the key worker [at nursery] and other kids better with the headphones".<br>"She continues to consistently wear the headphones at nursery for three days. The staff's view is that they make a massive difference there as I previously mentioned: more attentive and responsive to what staff say".<br>**Child**<br>"I know how to mute it. I was the person who showed my teacher how to do it, [before] when she was talking to another teacher she would hold the mic away as far as she could stretch her arm." | **Child** "I keep them in my tray. Miss T charges them on her desk."<br>"Could you give information to the teachers to do something for deaf awareness week".<br>"Nursery slowly got used to it." | "The teachers are worried about Covid and won't swap the microphone between teachers so she can't hear when she has a replacement teacher once a week and can't hear in PE."<br>"The teachers are so busy and stressed. I don't want to stress them more".<br>"The teacher is going to see how he goes and then decide whether we should use it" |
| **Impact on home life** | "The TV volume stays down now, we just place the microphone by the TV"<br>"The interaction between the boys [siblings] has been better"<br>"She [10y] is relying on it enormously..has paired it to everything in the house. It is now the way she watches TV. She has taken it out with her friends".<br>"She is getting on really well with the headphones - we've not tried them in school with the mic yet, however pairing them with the iPad at home is simply brilliant. She loves the freedom they give her and obviously, the hearing ability."<br>"There have been fewer arguments because he could communicate more effectively".<br>"Everyone at home raised their voices less" | His brother was fed up because he'd had the same trouble, he was like "why didn't I get anything like that?" | "If I am wearing the microphone and walk outside her bedroom the connection is often bad."<br>"I think a hearing aid would be easier so he could hear all the time"<br>"She doesn't want to wear it at home having worn it all day at nursery". |
| **Impact on fatigue** | "She is not as tired when she comes home from school"<br>"He is less tired after a day of wearing the headphones". |  |  |
| **Self confidence** | "His personality has come on a lot" | "I wasn't sure how she would cope with the other children [at school] but she wants to hear". |  |
| **Impact on the parent** | "If we hadn't been part of the study..I do worry about hearing..it's made me [parent] more chilled".<br>"My biggest worry was that she couldn't hear and her pronunciation sounds like a deaf child. Having the headphones, I'm not panicking that she is going back to where she was".<br>"I could talk to her at a normal level, rather than shouting."<br>"It makes me quite emotional how crazy she became when she tried them the first day...makes me realise that she could hear much better I think". | "Shopping trips were easier [when wearing the headphones], she could chat to me and she wasn't frightened she would lose me".<br>"He heard my voice, he smiled so much". |  |
| **Attention and concentration** | "He is very focused when wears the headphones. Beforehand it would have been 'what's that? Huh?'<br>"He has always struggled with concentrating and listening. His teacher says he looks up more [when wearing the headphones]."<br>"He'll listen to audiobooks to the end now rather than listening to only one or two pages" |  |  |
| **The headset** | "When I wore them at school my friends pretended to play guitar. My friends said 'when you were wearing the headset you look like a rock girl" ' "<br>"He thinks it's pretty cool, he thinks it's like a robot like thing".<br>"He is very protective over his headphones. The teacher was worrying about them getting damaged and I said 'no fear, he won't let anyone touch them'"<br>"The headphones give a visual cue to others that she has a hearing loss" | "Can children get their own from a shop? My friend wants one".<br>**Child** "I might need to wear them until I'm an adult. I might need a bigger size". | "A boy commented [teased] and now she really doesn't want to wear them at school."<br>**Child** "Tell other children to be careful with it, don't wear it at break times or lunch time. I am not wearing it in the playground" |
| **Social communication** | "Staff also comment that it makes a massive difference with friends - she can hear and interact with them better and they can understand what she says" | "When you are learning language, life, and social skills, whether it is small or not, you are missing something [if you can't hear]" |  |
| **The Hear Glue Ear app** | “The app appeals to her” |  | “The app needs to have an older children section”.<br>“she has got through all of the stories songs and games on the app [in lockdown], the app needs to have more on it.” |
| **General comments** | “I can’t tell you how much it has helped”.<br>“It has made a massive difference to her.”<br>“It’s nice to get something to help really”.<br>“This has really helped” .<br>“All his violin lessons for 10 months have been online. He used the headphones paired to iPad for zoom lessons. He just passed his exam”.<br>“We were so desperate and we had run out of options. This is just brilliant. We are over the moon to be part of the study”.<br>“I have been wanting to support [my child] with something like this”.<br>“We are grateful for them...the first two years at school are so important.”<br>“He was excited to have something that was just his”. | “We would rather avoid grommets”.<br>“We were quite shocked at how low her hearing was when tested last year and the need for her to have an operation, personally I’d like to avoid that if possible, but just don’t want her to feel isolated and fall behind.” | “If she had only had this a few years ago. We needed it then.”<br>“I wish we had been given this while he was at primary school. We didn’t think it was a good idea for him to wear them as he starts secondary school” |

## Appendix 2. Instructions for parents and teachers

### Hear Glue Ear Assistive Hearing Set for children: Instructions for Use

#### What is the Hear Glue Ear assistive hearing set?

This set comprises two devices: a bone conduction headset, and a microphone. There is a video about setting up this product on website www.hearglueear.wordpress.com the ‘menu’ has an option at the bottom called “Research during covid-19” which has all the information about this study and videos for use.

The headset uses vibrations to enable your child to hear sound better. The vibrations go through their cheek bone and into their inner ear, avoiding the middle ear. This will assist your child to hear more clearly if they have a conductive hearing loss.

The microphone is used to transmit sound to the headset.

The headset can also be used like a set of normal headphones when paired with a mobile device (such as a tablet or mobile phone) to allow your child to hear high quality sounds when watching programmes, listening to audiobooks or playing games which are all available on the free Hear Glue Ear app which we would like you to download from the App Store. The Hear Glue Ear app logo looks like this.

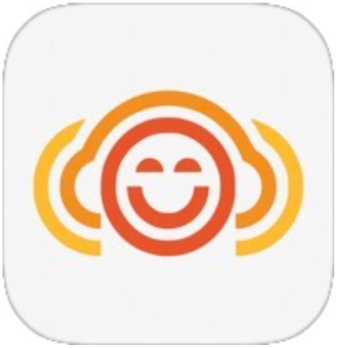

For reassurance the Hear Glue Ear app is registered as a class 1 medical device and has been approved and recommended by professional audiology and medical bodies such as the, Organisation for the Review of Care and Health Apps, the British Audiology Association, and the National Institute for Health and Care excellence. It also won Children’s app of the year at the UK app awards in 2019.

**Figure 1:**
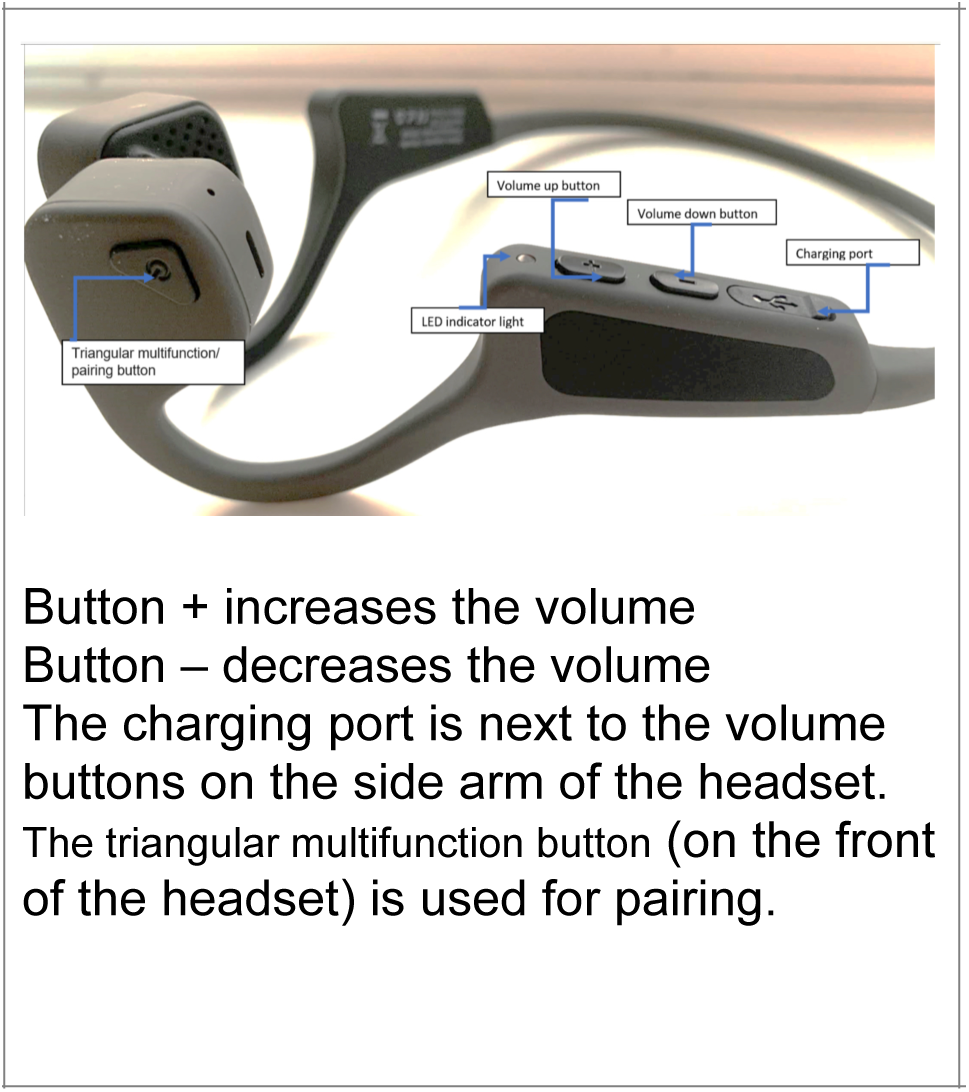
The Headset.

**Figure 2:**
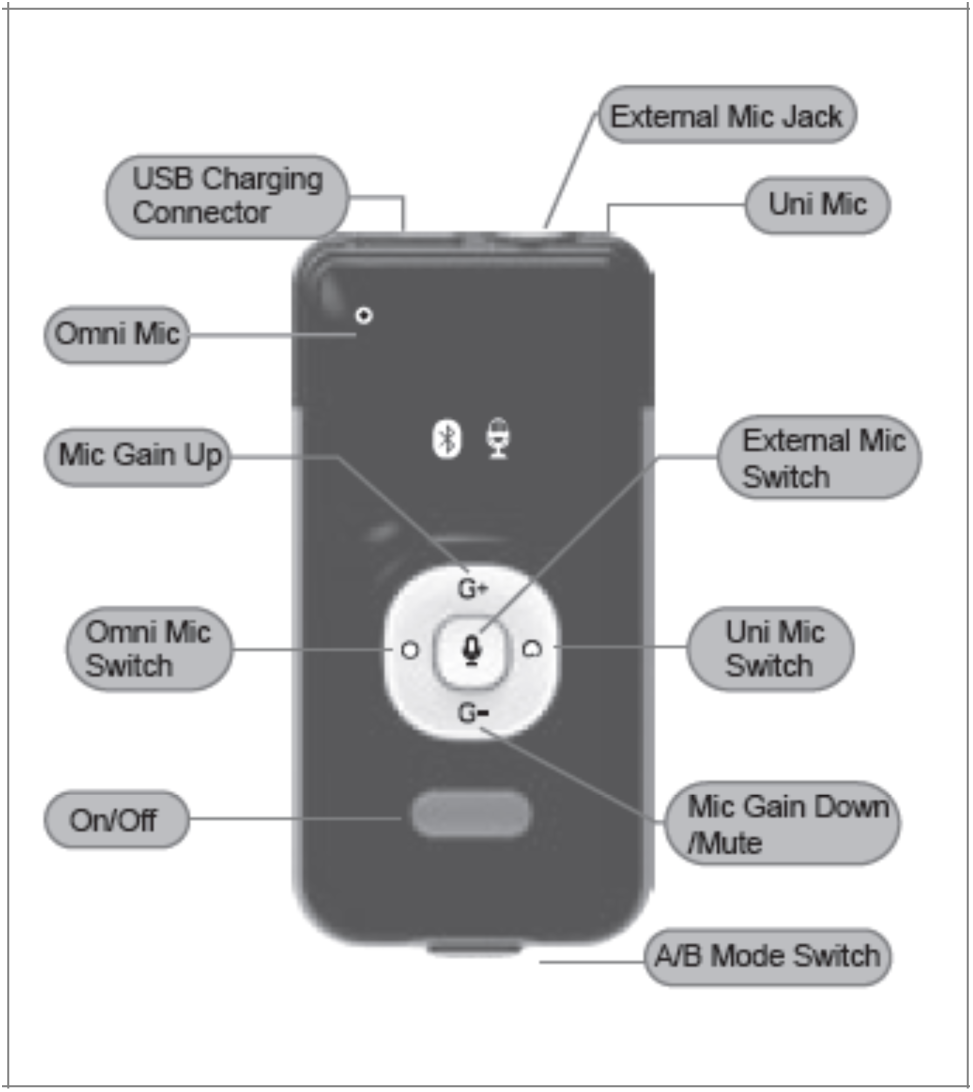
The Microphone.

Your child can wear the headset at school, at home or for speech and language therapy. The microphone should be worn by the person whose voice your child needs to hear clearly, such as a teacher, parent, therapist or other family member/friend. An introduction sheet for teachers can be found at the bottom of this instruction sheet.

The headset can be worn during sports such as PE, running, cycling etc, but cannot be worn while swimming or bathing.

### Intended use

This product set is intended to be used to aid hearing in children diagnosed with a mild – moderate conductive hearing loss. This conductive hearing loss could be caused by Glue Ear (a build-up of fluid/mucus behind the ear drum) or other causes. The product set is not indicated for children with purely sensorineural hearing loss. Your research team or clinical team will prescribe use of this product only if your child has the correct type of hearing loss. The products can be used for up to 4 -6 hours per day before needing to be recharged. It is recommended that they are used at school, in situations where there is background noise or, if your child is struggling to hear instructions. Your child should be encouraged to make adults aware if the sound is too loud or if they can’t hear so that volume levels can be adjusted. They can always take the headset off, if they would like a break from wearing it. Please see the section of volume levels for adjusting if needed.

#### Instructions for Use

##### Using the product set

Before use, please check that the product has all the correct parts included:

- The headset
- Charging cable for headset
- The microphone
- Charging cable for microphone
- Instructions for use

###### Charging

Both elements of the system, the headset and the microphone, need to be fully charged prior to use.

The leads can be connected to a USB port on a computer or laptop, or to a USB plug for charging, as shown.

When the microphone is charging, the amber LED will light up. It will turn off when the microphone is fully charged.

The headset takes an hour and a half to completely charge. When charging the LED will be red. Once charged the LED will turn blue.

WARNING: Only use the provided leads to charge the devices. WARNING: Do not use the devices whilst charging.

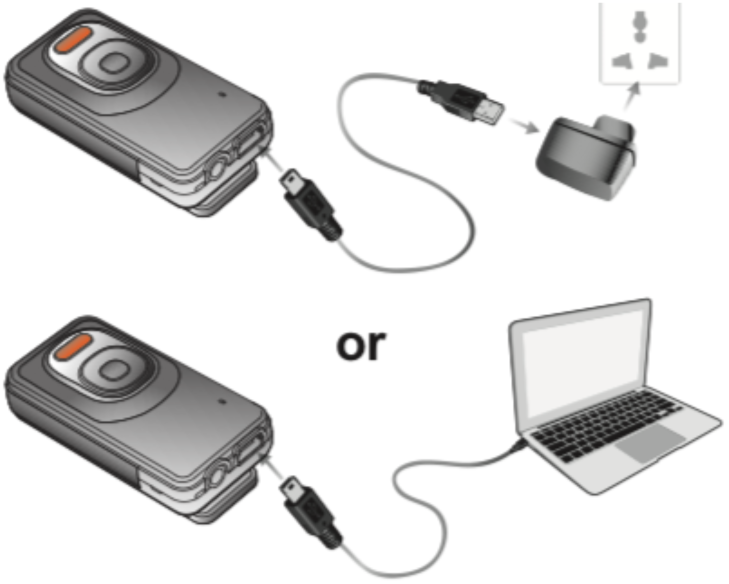

###### Wearing the headset and microphone

Videos are available to follow on the www.hearglueear.wordpress.com website under the section ‘Research during Covid-19’. Your clinical team will also be able to tell or show you how to wear the headset during remote clinical consultation. During use, the headset is worn such that the vibrating speakers sit just in front of the ears, on the cheek bones. The headband sits behind the neck, as seen in Figure 4a.

To check if the headset is working, an adult can place the headset on their cheek bones as shown in Figure 4b.

The microphone needs to be worn securely attached to clothing, with no lanyards or scarves worn around the neck, as shown in Figure 4c. Adjust the direction of the microphone to your mouth by rotating the Bluetooth Microphone.

###### Using your headset with the microphone

1. Turn on the headset (it will say “power on”). To pair the headset with any device HOLD your finger down on the triangular multifunction button (see picture below) for 5 seconds, until the LED flashes blue and red and the device says “Pairing”.

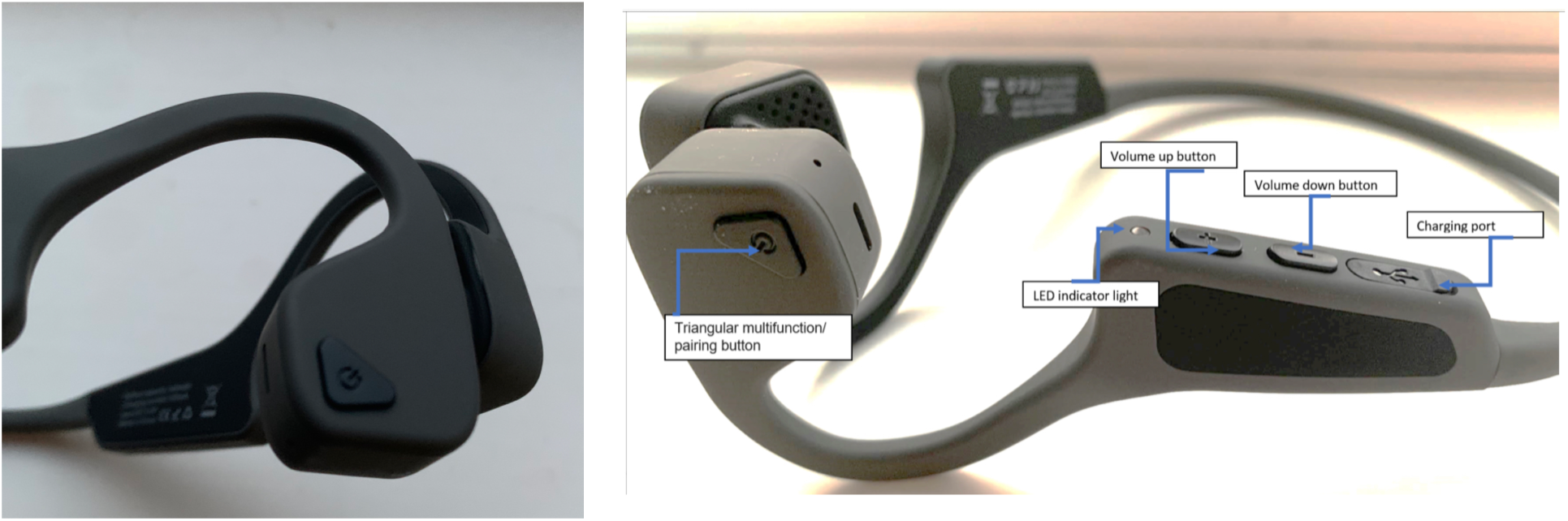
2. Ensure that the microphone is switched to Type A:

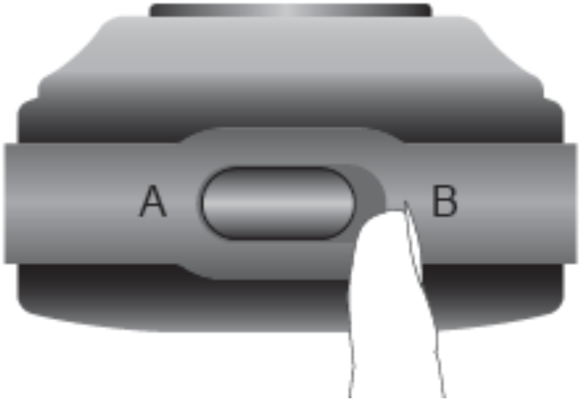
3. Turn on the microphone on and into Type A pairing by pressing the on/off button for 2 seconds until the LED flashes blue

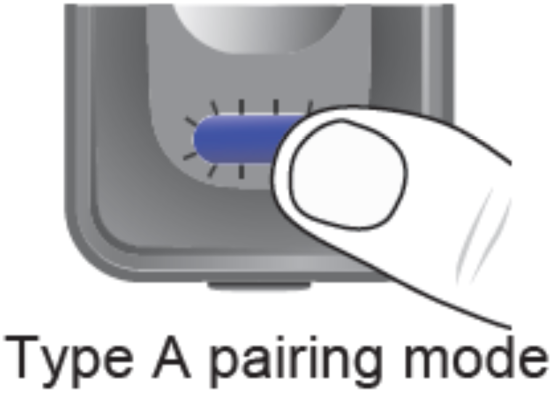
4. The headset will automatically connect to the microphone and say “Connected”. The blue LED on the microphone will change to flash slowly. (This can take up to 30 seconds).
5. The headset can now be placed on your child’s head as shown in Figure 4a.
6. Remember to turn both the microphone and headset off after use: To turn off the microphone, press and hold the on/off button for 2 seconds, the amber LED will stay on for 1 second and then turn off. Release the on/off button. To turn off the headset, press and hold the on/off button. You will hear the headset say, “power off”.

###### Using the Microphone

The microphone can be used in unidirectional and omnidirectional settings. Both have their advantages for different applications. It is important to make sure that you are speaking into the correct mic depending on the setting:

If you wish to use the external microphone attach this to the audio port and select the External Mic button on the microphone:

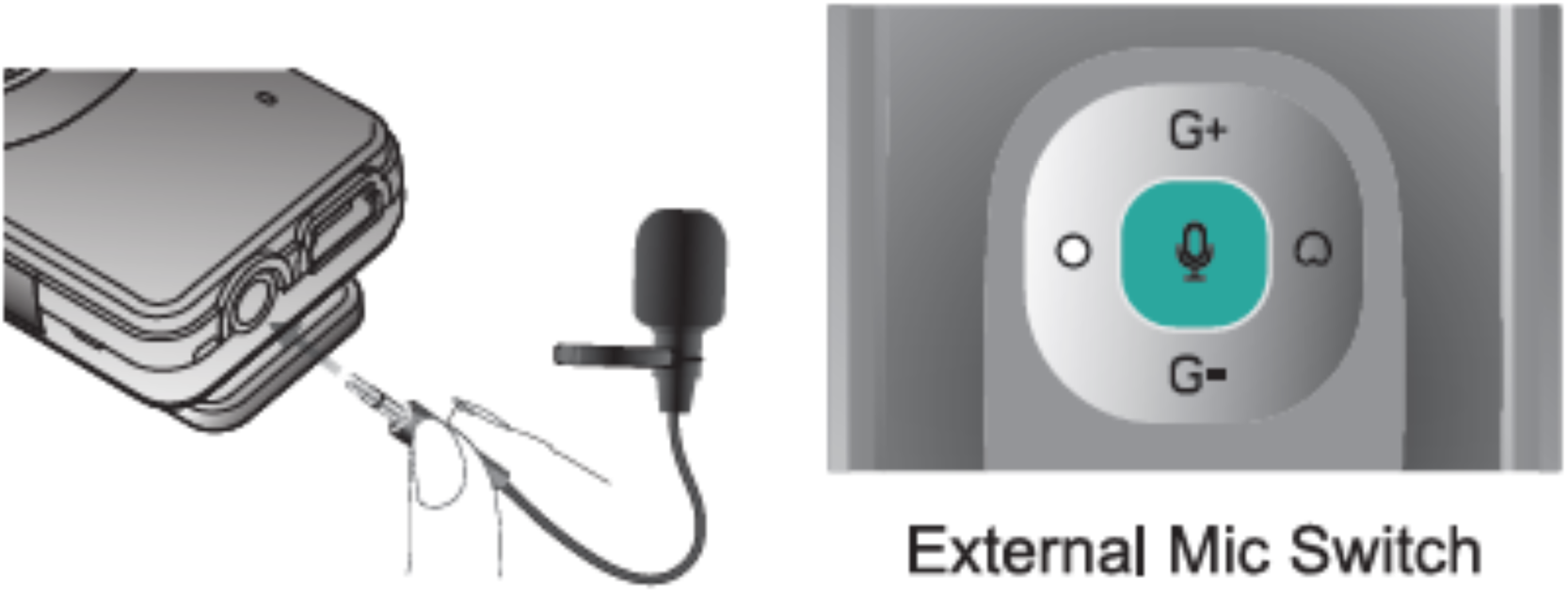

If you wish to mute the microphone, press and hold the G-button for 2 seconds. Press the G+ button to unmute. Whilst the microphone is muted, the power button LED will flash blue-amber-blue.

###### Using your headset with smart phone or tablet

You will need to pair the headset to your smart phone or tablet to use the Hear Glue Ear app. Pairing enables the two devices to talk to each other.

1. Ensure that the microphone is turned off. You will not need the microphone for this.
2. Turn on Bluetooth on your mobile device
3. Press and hold the on/off button on the headset for 5 seconds until the headset says “Pairing” and the LED blinks blue and red.
4. The headset should now be visible on your mobile device as “G18”. Select the headset on your mobile device and the two products should then pair.
5. The headset will say “Connected” once the pairing is complete.
6. Once paired, any sound from the mobile device will be sent to the headset.

###### Warning

Be aware that all sound from the mobile device will be transmitted to the headset. Remember to disconnect or turn off Bluetooth once the child has finished using the mobile device.

###### Volume levels

The volume level will have been set at a default mid volume setting. Always adjust the volume from the headset: never use the microphone to adjust the volume. If your child complains about the volume level, you are able to adjust this by using Button + on the headset to increase volume, and Button - to decrease volume. This must be done whilst sound is playing through the headset.

We advise to only change the volume by one or two increments each way. The base volume level set is at the middle of the volume range of the device. If you need to reset to this level, please follow the procedure below:

1. Pair headset to a mobile device and play music through the mobile device.
2. Using ‘Button -’ to turn the volume down as low as possible. You will hear a tone each time you press the button. At the bottom of the volume level you will hear another tone.
3. From the lowest level, use ‘Button +’ to increase the volume 8 times. This is the middle of the volume range.

###### Trouble shooting

You can check if the device is functioning correctly by placing the headset on to your own cheek bones as shown in Figure 4b and tapping or blowing on the microphone. You should hear the sounds through the headphones.

###### Lights and indications on the product

| Indication | Meaning | Action Required |
| --- | --- | --- |
| Headset |  |  |
| LED Solid red colour | Charging | None |
| LED Solid blue colour | Charging complete | None |
| LED Flashes red and blue | Pairing mode | Pair to microphone or mobile device |
| LED Blue double flash every 3 seconds | Connected | Connected to phone tablet or microphone |
| LED rapidly flashes red | Low battery | Charge device |
| Microphone |  |  |
| LED amber colour | Charging | None |
| LED flashes blue every two seconds | Paired to headset | None |
| LED flashes blue every five seconds | Not connected | Turn both headset and microphone off and follow Instructions to pair to headset |
| LED flashes blue rapidly | Type A pairing mode | Ensure headset is in pairing mode |
| LED flashes blue and amber alternately | Type B pairing mode | Switch to type A pairing mode and follow instructions to pair to headset |
| LED flashes blue-amber-blue | Mute | Press the G+ button to unmute |

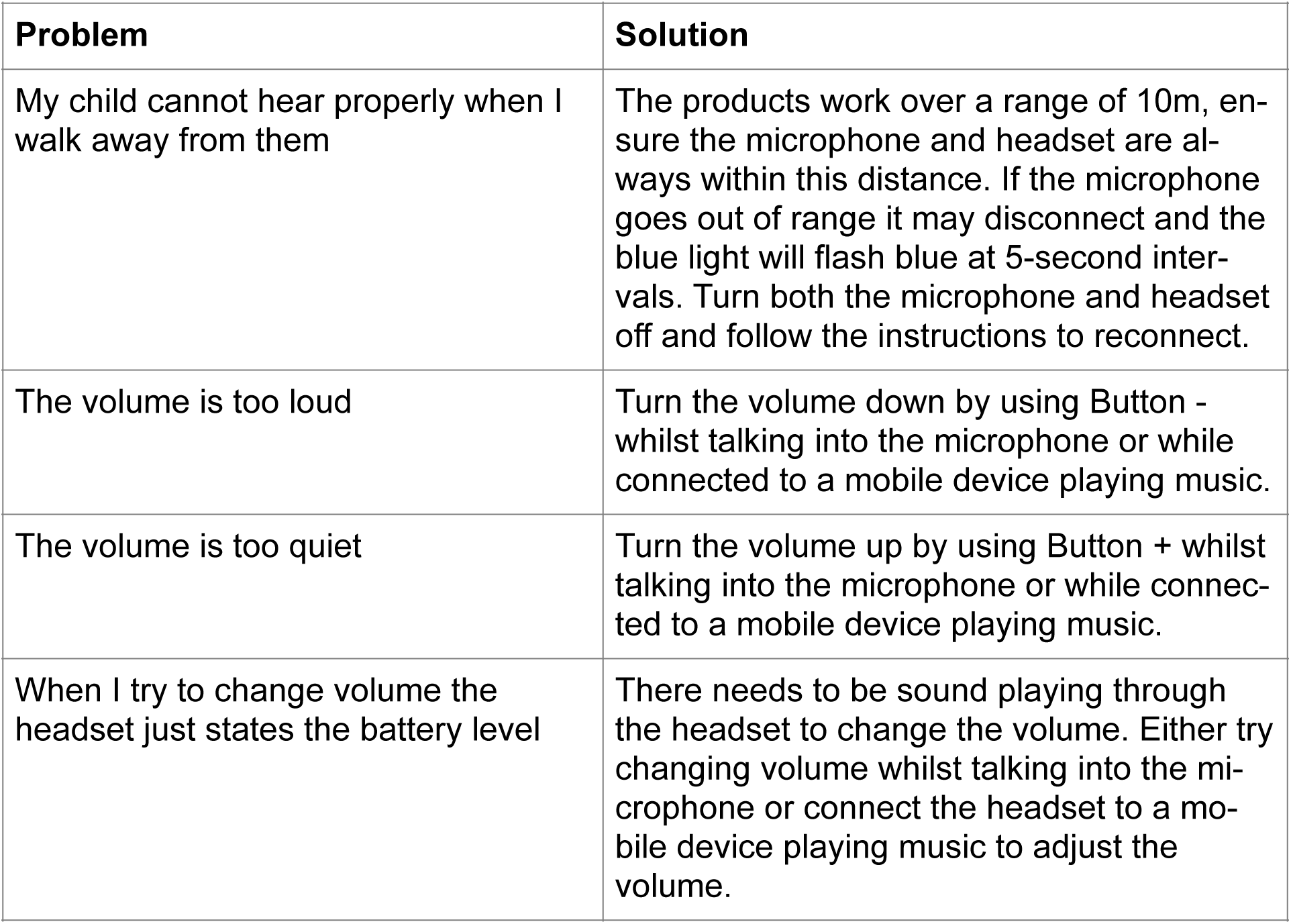
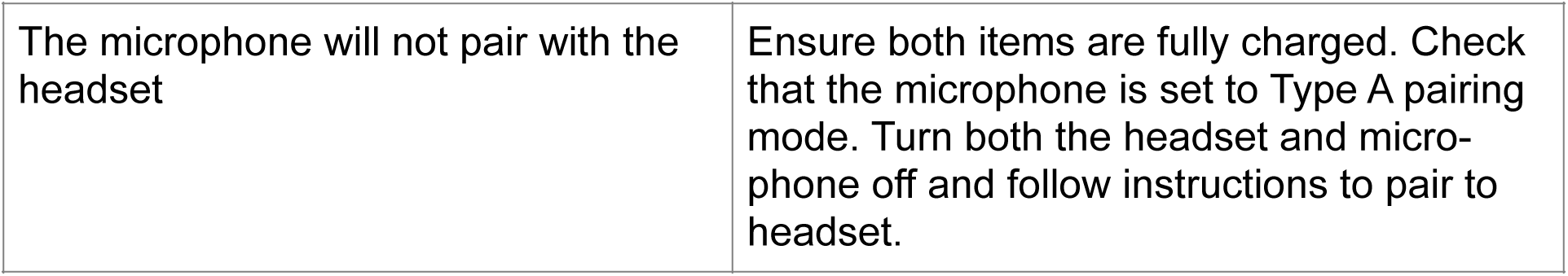

**Warning**: Neither product contains user serviceable parts. Do not attempt to dismantle or open the products.

**Warning**: The product cannot be used while it is charging.

**Warning**: This product cannot be used under water. It is water resistant but not waterproof. Do not wear for swimming, bathing or showering. If the product gets wet, turn it off and allow it to dry naturally before using.

###### Disposal

The products should not be disposed of in normal household waste. They must be returned to the clinical department who issued them.

### Information for Teachers

Thank you for agreeing to use the Hear Glue Ear Assistive Hearing Products in the classroom – here is our guide to activating the device to allow the child to hear well. If you have used an ‘FM’ device before, then this works in a very similar way.

#### The reason the child needs to use the device

The device is for children who cannot hear properly because the middle part of their ear does not work correctly. This might be a temporary or permanent problem. For the child it feels a bit like their ears are blocked, or as if they are walking around with their fingers in their ears all day. This headset directs sound along the child’s cheekbones, bypassing the middle part of the ear, and accessing the normal part of their inner hearing instead. Therefore, with the headset the child hears more easily. The child and their family has agreed to be part of a research study which is assessing children using this product when other services are reduced, closed or not easily accessed as a result of the coronavirus pandemic.

#### How to operate

1. Remove all staff badges, scarves or necklaces that may touch the microphone or interfere with it. Clip it onto your clothing.
2. Turn the headset on:
  a. EITHER …Ask the child to turn on their headset – the parent may have already done this. OR…
  b. If you need to help by turning the headset on for the child, the easiest thing to do, is to place the headset on your own cheekbones (as in the picture). Press and hold the on/off button (The triangular button on the front of the headset) for 5 seconds, until the LED flashes blue and red and the device says “Pairing”.

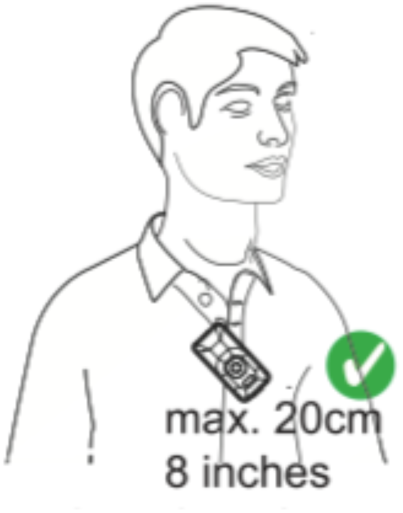
3. Ensure that the microphone is switched to Type A:

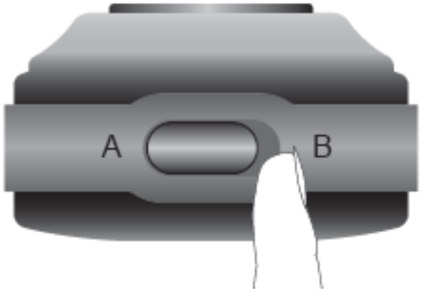
4. Turn on the microphone on and into Type A pairing by pressing the on/off button for 2 seconds until the LED flashes blue

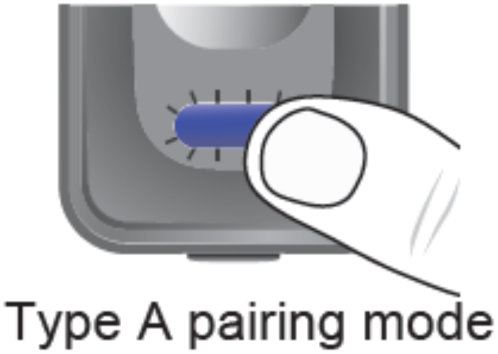
5. The headset will automatically connect to the microphone and say “Connected”. The blue LED on the microphone will change to flash slowly. (Do not be concerned if this takes up to 30 seconds).
6. The headset can now be placed on your child’s head as shown in Figure 4a.
7. Remember to turn both the microphone and headset off after use: To turn off the microphone, press and hold the on/off button for 2 seconds, the amber LED will stay on for 1 second and then turn off. Release the on/off button.

To turn off the headset, press and hold the on/off button.

To reduce background noise, use the microphone in unidirectional mode by pressing the button below:

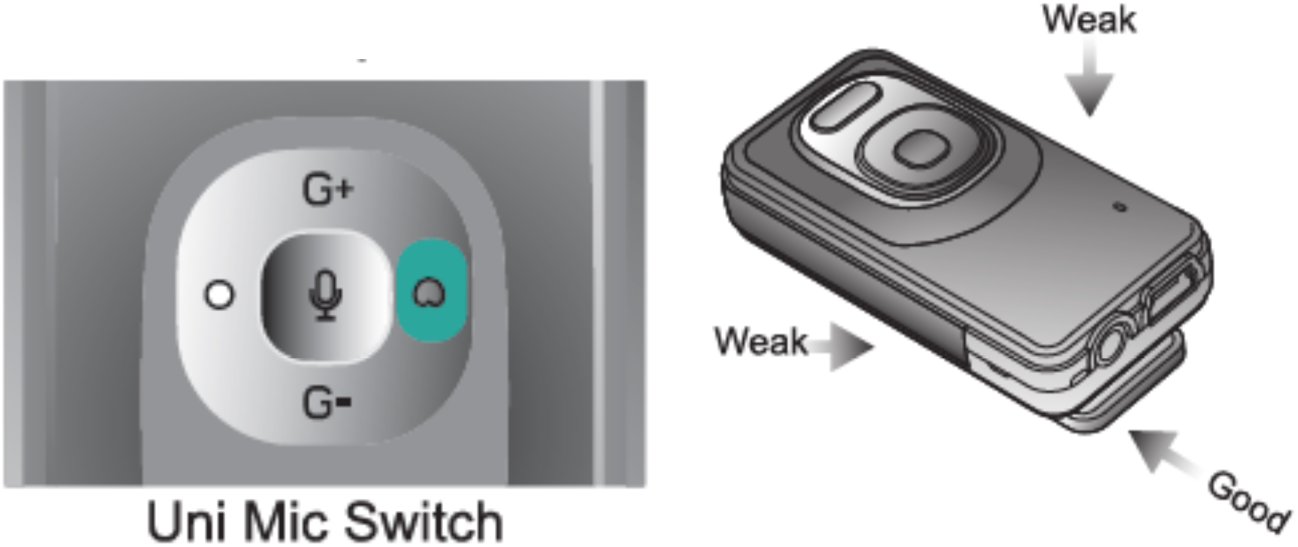

#### MUTING THE MICROPHONE

If you wish to mute the microphone (e.g. when speaking to other children) press and hold the G-button for 2 seconds. Press the G+ button to unmute. Whilst the microphone is muted, the power button LED will flash blue-amber-blue.

## Appendix 3

### HEALTH CARE PLAN FOR A CHILD/YOUNG PERSON WITH GLUE EAR (Template2020)

Glue ear can also be called Otitis Media with Effusion (OME)

#### PERSONAL DETAILS

|  |
| --- |
| Name of Child/Young Person: |
| Date of Birth: |
| Name of School/Setting: |
| NHS No: |
| Date Care Plan Completed: |
| Date Care Plan to be Reviewed: As care needs change |

#### CONTACT INFORMATION

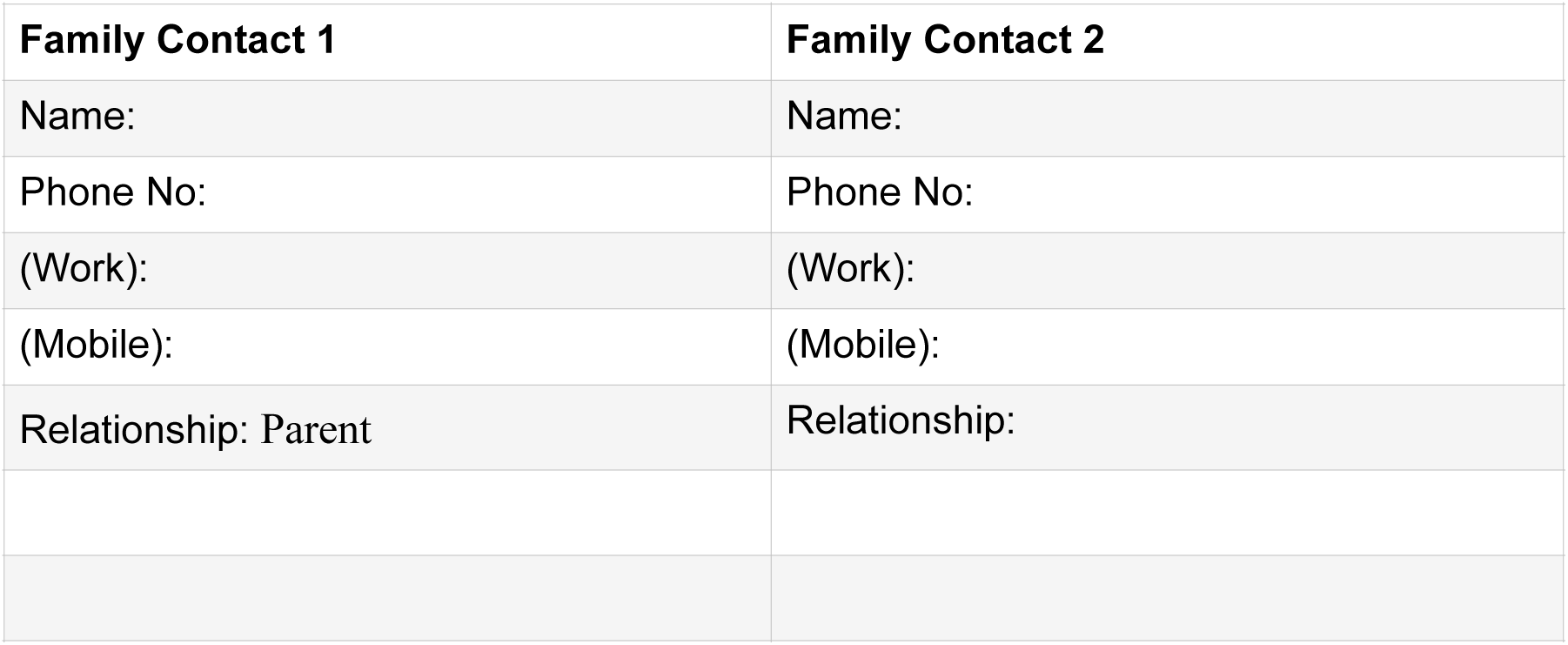

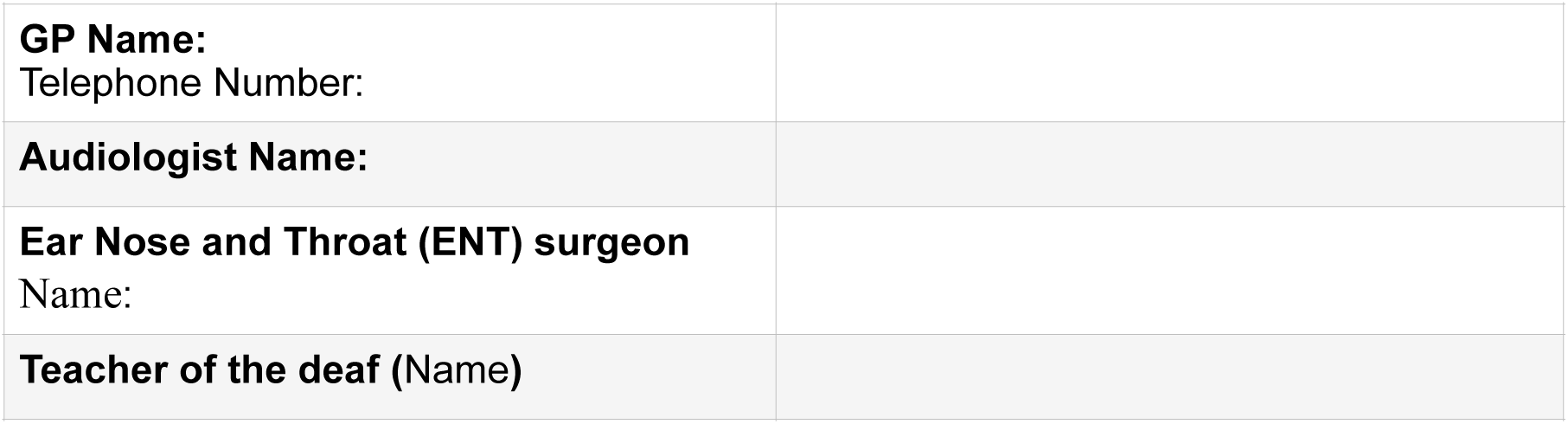

#### Note for parents

Parents/carers are reminded of the importance of informing schools or adults caring for their child of any changes in hearing support /surgery or ongoing concerns/changes in hearing health.

##### •CONFIDENTIALITY

For reasons of safety and rapid access, this form may be displayed on a notice board in the staff room.

##### Copies held by

Parents/Community records/School/Consultant/Audiologist/ENT surgeon/ Other*

*Delete as applicable

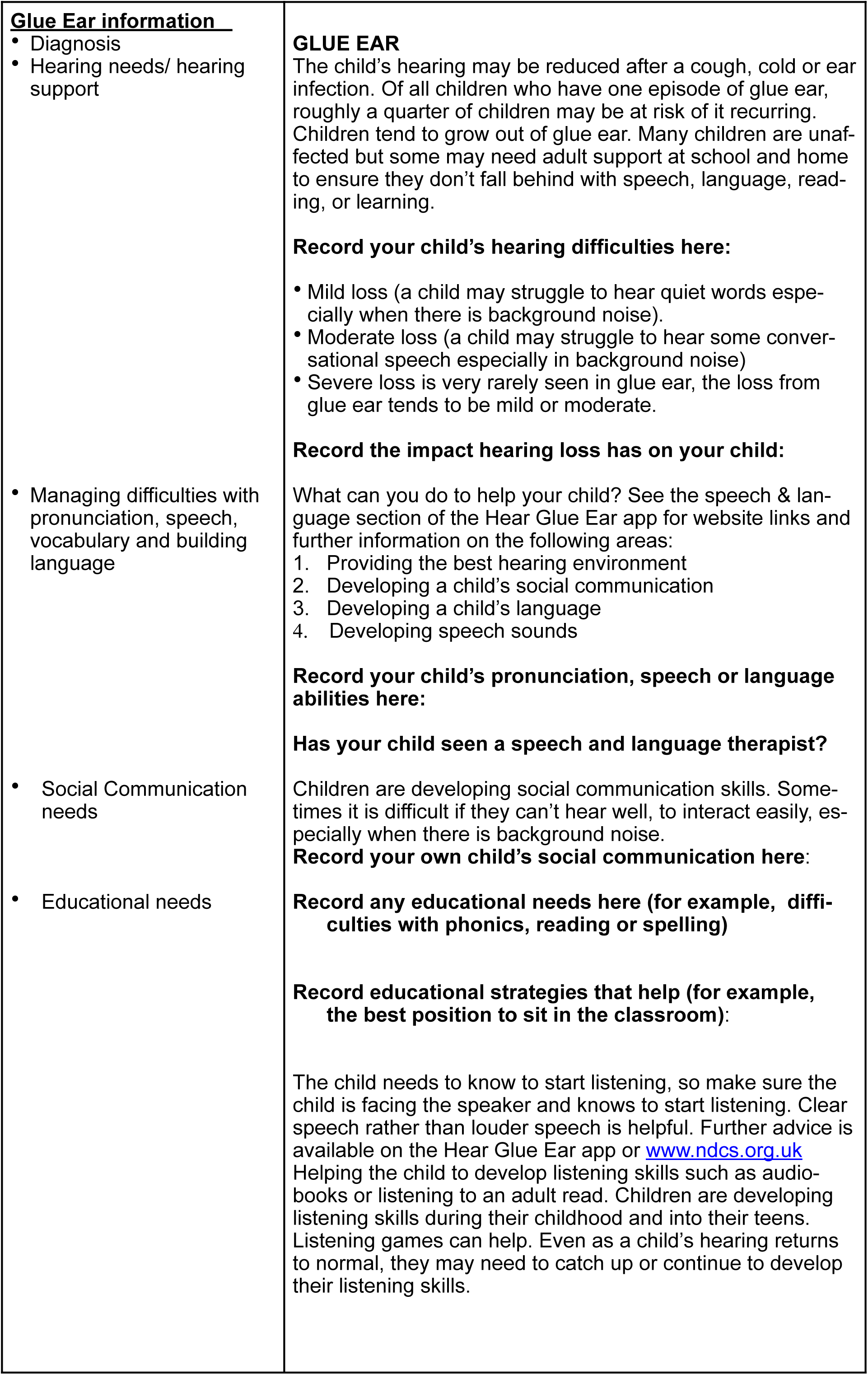
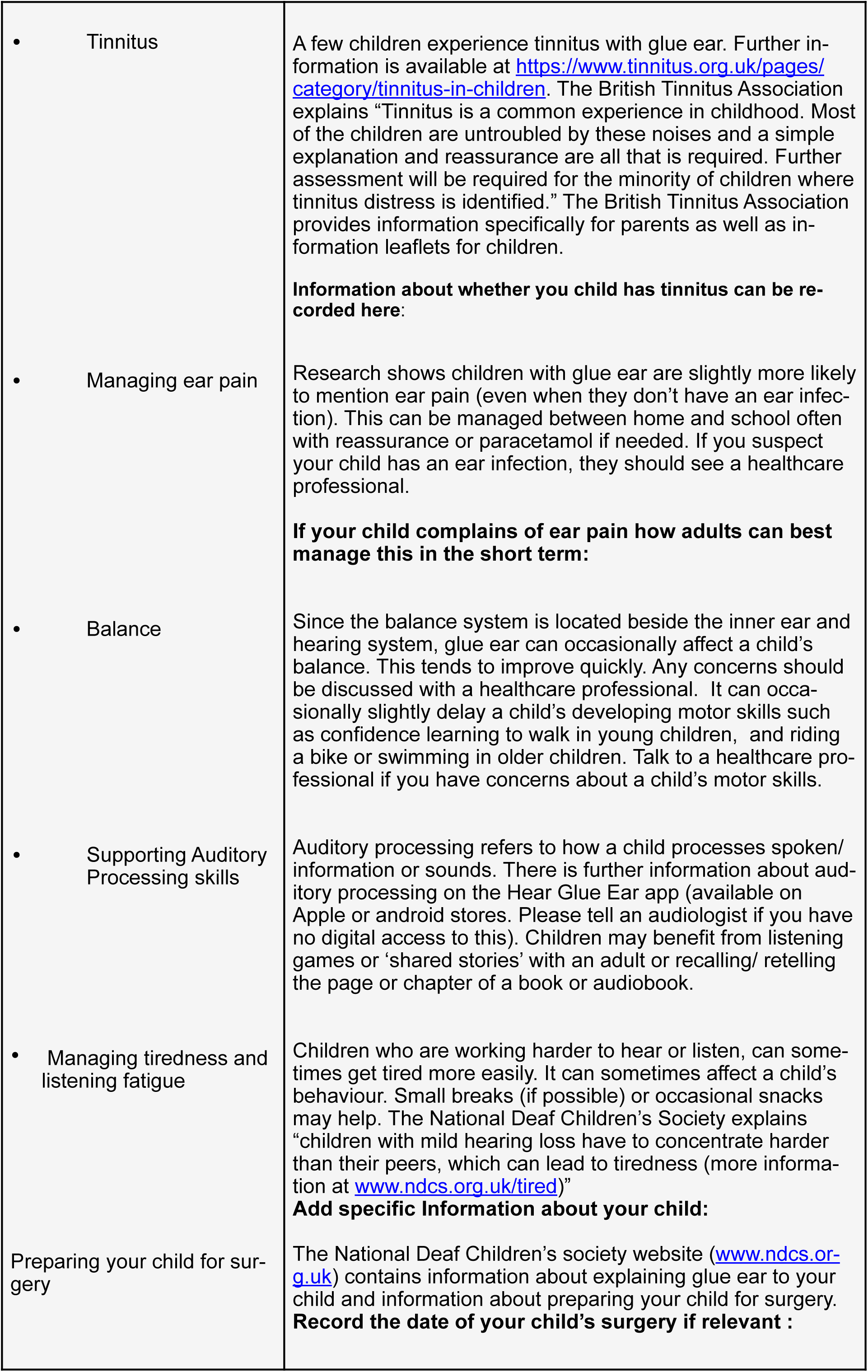

#### Equipment

**Please record here if your child is using special equipment or aids in school or at home including use of Hear Glue Ear app or type of headphones used:**

|  |  |  |
| --- | --- | --- |
| **1. When should the child be referred for a hearing test?** |  |  |
| • If hearing loss lasts longer than a few weeks after a cough or cold parents may want to discuss with a healthcare professional whether a referral for another hearing test is needed.<br>• If a child has ongoing speech, language or reading concerns, a speech and language therapist may suggest referral for a hearing test. |  |  |
| **2. What additional support might the child need?** |  |  |
| • Many children with glue ear do not fall behind at school, but a few may need adult support to catch up with listening skills, reading, phonics or spelling at home/school after an episode of glue ear. The child may need to be referred for another hearing test if glue ear is suspected again. |  |  |
| **3. Other considerations:** |  |  |
| Swimming: This depends on ENT advice given particularly after a grommet operation. If the ear is to be kept dry, swim ear moulds, or over-the-counter ear plugs can be purchased, some children also wear swim ear bands to ensure the ear moulds or ear plugs stay in place.<br>Physical Education: All usual care (note that balance concerns can sometimes affect motor skills. Usually, the benefits of maintaining normal physical activity exceed the risks of injury due to balance problems)<br>Other concerns: (record here) |  |  |
| **<u>Health Care Plan Agreed by:</u>** |  |  |
| Parent/ Guardian of child | Signature | Date |
| Head teacher/ head of receiving organisation: | Signature | Date |
| Healthcare professional / Other | Signature | Date |

### Glue Ear Passport

**(to share between health professionals and school. Recommend a copy to be kept in school staff room) *Date***

**Hear Glue Ear (Supporting child/ young person with glue ear)**

**Name: Year: Date of birth: Teacher of the Deaf:**

C**ontact:**

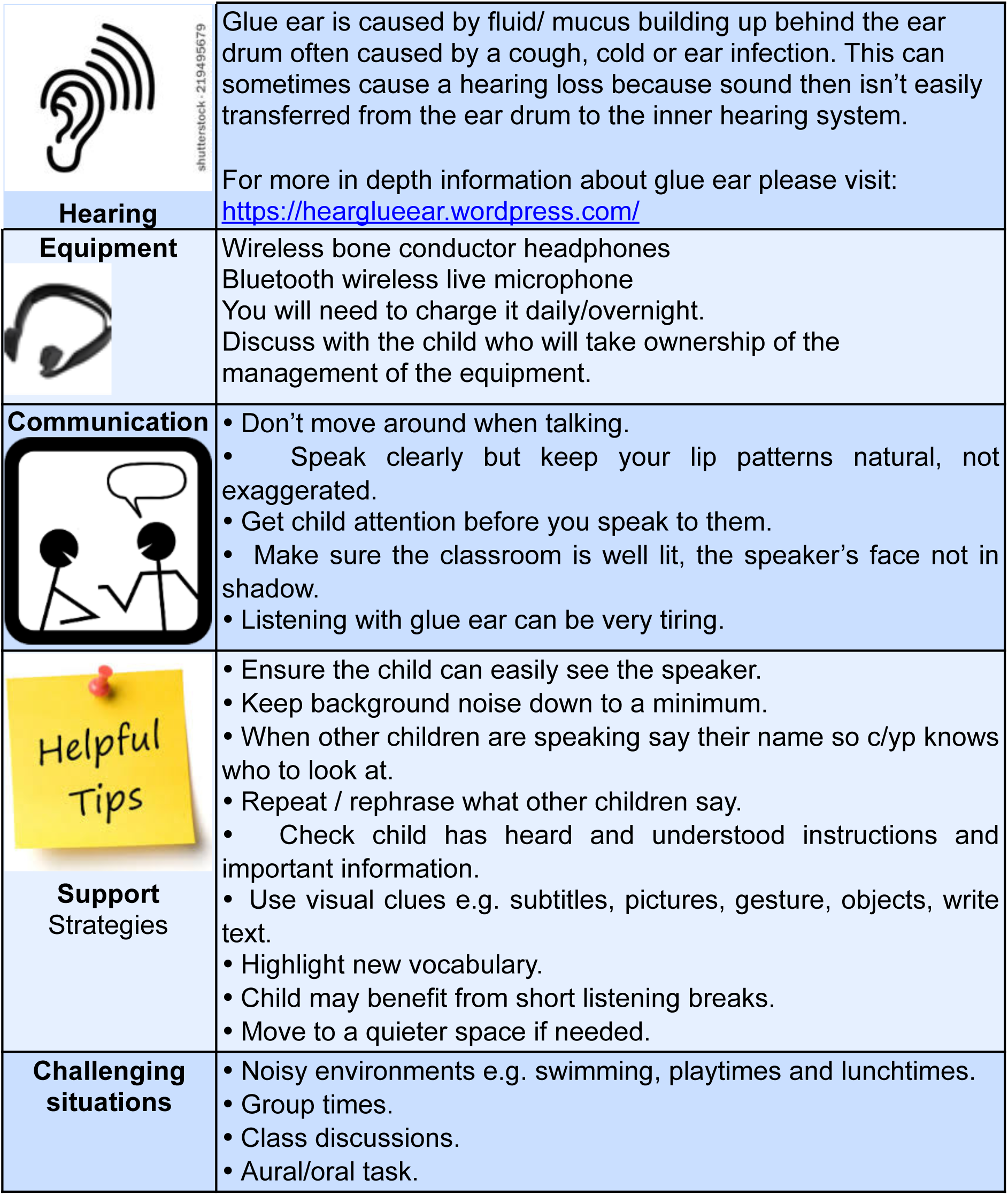

### Author declaration of interests

Dr Tamsin Holland Brown conceptualised the use of the kit and helped create the free, charity funded, Hear Glue Ear app that was used in this study. Dr Holland Brown has not received funding or payment related to the Hear Glue Ear app.

None of the authors have received any funding or payment for completing the research nor benefitted from sales of the equipment.

Other authors declare no conflict of interest.

### Protocol

Details of this study including the protocol are available on the following link: https://hearglueear.wordpress.com/research-during-covid-for-children-with-glue-ear-who-have-been-considered-for-a-grommet-operation/

### Data sharing

of anonymised patient data collected for the study can be made available to others. Other details of the study have been published on https://hearglueear.wordpress.com/

### Sources of Support and acknowledgments

The authors would like to thank Dr Josephine Marriage (CEO of Chear) for helping with planning of the study, Dr Alison Sansome (Consultant Paediatrician, Cambridgeshire Community Services NHS Trust) and Dr Jacqueline Taylor (Consultant Paediatrician, Cambridgeshire Community Services NHS Trust) with supporting the study to run during COVID-19 despite disruption to the department. Also the Department of ENT surgeons at Cambridge University Hospitals NHS Foundation Trust) for their support, consecutive leads Mr Roger Gray and Mr James Tysome, with Mr Nico Jonas and Neil Donnelly as paediatric otology and MDT leads. Thank you to Ivy Court for discussions and Cambridge Hearing Trust charity who funded the Hear Glue Ear app and Cambridge Digital Health who created the Hear Glue Ear app. Improvements made to the Hear Glue Ear app early in COVID-19 were funded by NHS Charities and Cambridgeshire Community Foundation. The Perivoli Foundation provided funding for equipment to be accessible to services leading on from this research. Thank you to sensory support teachers Sibel Djemal, Amarinda Benson and Angela Howgate for the glue ear passport and moving the project forwards. Finally thank you to children Daisy, Josca, Sunny, Lilac and Marigold for their patience and joy while their parents covered services during COVID-19 as well Mum working on this research project.

